# Celecoxib Colorectal Bioavailability and Chemopreventive Response in Familial Adenomatous Polyposis Patients

**DOI:** 10.1101/2020.12.02.20242214

**Authors:** Peiying Yang, Xiangsheng Zuo, Shailesh Advani, Bo Wei, Jessica Malek, R. Sue Day, Imad Shureiqi

**Affiliations:** Departments of Palliative, Rehabilitation, and Integrative Medicine, The University of Texas MD Anderson Cancer Center, Houston, Texas; Departments of Clinical Cancer Prevention, The University of Texas MD Anderson Cancer Center, Houston, Texas; Departments of Gastrointestinal Medical Oncology, The University of Texas MD Anderson Cancer Center, Houston, Texas; Department of Oncology, Lombardi Comprehensive Cancer Center, Georgetown University, Washington DC; Department of Epidemiology, School of Public Health, The University of Texas Health Science Center, Houston, Texas

**Keywords:** celecoxib, polyps, colorectal cancer, chemoprevention, lipoxygenase

## Abstract

The chemopreventive activity of celecoxib against colorectal cancer is limited to a proportion of familial adenomatous polyposis (FAP) patients who experience a response. The cause of this response variability and the potential mechanisms underlying these responses remain poorly understood. Preclinical studies showed that celecoxib increases the production of main oxidative metabolism product of linoleic acid, 13-S-hydroxyoctadecadienoic acid (13-S-HODE), to suppress colorectal tumorigenesis. We conducted a phase II clinical study to determine whether celecoxib increases 13-S-HODE production in colonic adenomas from FAP patients. Twenty seven FAP patients completed a 6-month oral course of 400 mg of celecoxib twice a day and had colonoscopies before and after celecoxib treatment to assess colorectal polyp tumor burden and obtain colorectal normal and polyp biopsies to measure celecoxib, 13-HODE, 15-HETE, 12-HETE, and LTB4 levels by LC/MS. Celecoxib levels in sera from those patients were also measured before treatment and 2, 4, and 6 months of treatment. Seventeen of the 27 patients experienced a response to celecoxib, with a more than 30% reduction of colonic polyp burden on the basis of a reproducible quantitative assessment of colonoscopy results. Celecoxib levels were significantly lower in polyp tissues than in normal colorectal tissues. Celecoxib levels in sera and normal colorectal tissues were correlated in patients who experienced a response to celecoxib but not in those who did not. Among the measured lipoxygenase products, only 13-HODE levels were significantly lower in polyp tissues than in normal tissues. Our findings demonstrate the differential bioavailability of celecoxib between normal and polyp tissues and its potential effects on clinical response in FAP patients.

## Introduction

Colorectal cancer (CRC) is the third leading cause of cancer death in the United States (1). Chemoprevention is one promising approach to reduce the CRC health burden; it has been found to be feasible in preclinical and early clinical studies (2). Non-steroidal anti-inflammatory drugs (NSAIDs) are a very promising class of chemopreventive agents in CRC, as shown in epidemiologic, preclinical, and clinical studies (3). Nevertheless, the currently estimated efficacy rate of NSAIDs in CRC chemoprevention is 50% or less (2-5). Furthermore, NSAIDs, especially COX-2 inhibitors such as celecoxib, have been associated with an increased risk of cardiovascular events (6) (7). Thus, identifying patients who might benefit from this chemopreventive approach is imperative to better tailor treatment to maximize the benefit versus the risk.

Familial adenomatous polyposis (FAP) is an autosomal dominant hereditary cancer syndrome with 100% penetrance; it is characterized by the early onset of a large number of colorectal adenomas that invariably lead to CRC development before the age of 40 years (8). Thus, the development of effective chemopreventive interventions for these patients is important. In FAP clinical trials, celecoxib reduced the colorectal adenoma burden in approximately 50% of patients (9). NSAIDs were initially believed to act by inhibiting prostaglandin synthesis; however, various reports have questioned this concept, including one that showed that PGE2 inhibition is not correlated with the reduction in polyp burden by celecoxib in FAP patients (10). Thus, more predictive mechanistic biomarkers of NSAIDs’ chemopreventive effects are still needed.

Lipoxygenases (LOXs) are a class of enzymes that are involved in polyunsaturated fatty acid metabolism. Within this family, 15-lipoxygenase-1 (15-LOX-1) is the main enzyme that metabolizes linoleic acid, the predominant polyunsaturated fatty acid in the human diet, to produce 13-*S*-hydroxyoctadecadienoic acid (13-*S*-HODE), which has differential anti-tumorigenic effects compared to those of other LOX products (11). 15-LOX-1 is downregulated in up to 100% of human-invasive CRC (12) and in colorectal adenomas in FAP and non-FAP patients (12-15). The product profile of LOX in FAP and sporadic colorectal adenoma patients revealed that 13-HODE levels were lower in polyps than in paired normal colorectal tissues; this was the only significant alteration (13).

NSAIDs’ induce apoptosis in CRC cells, which plays an essential role in their chemoprotective effects (16,17). Celecoxib upregulates 15-LOX-1 expression and increases 13-*S*-HODE production during apoptosis induction; these events were found to be critical to celecoxib’s ability to induce apoptosis in preclinical models (18). However, whether celecoxib modulates LOX metabolism to exert chemopreventive effects clinically remains unclear. The current study examined the clinical relevance of celecoxib modulation of metabolism via LOXs to its chemopreventive effects in colonic adenomas of FAP patients.

## METHODS

### Clinical study and human colonic tissue sampling

The Institutional Review Board of The University of Texas MD Anderson Cancer Center (Houston, Texas) approved our study. All patients gave informed consent before participating. This single-arm celecoxib study enrolled 47 FAP patients at The University of Texas MD Anderson Cancer Center between November 2004 and May 2010. Twenty-seven patients completed the procedures required for the reported laboratory analyses. A baseline colonoscopy (or sigmoidoscopy in patients who had undergone colectomy) was performed before the initiation of celecoxib, and a follow-up colonoscopy or sigmoidoscopy was performed after celecoxib treatment. The celecoxib dose was 400 mg orally twice daily for 6 months. Patients’ demographic and clinical characteristics, treatment schedules and polyp burden assessment to determine response have been described previously (19). Celecoxib levels in sera were measured and assessed prior to initiation of the study and then at 2, 4, and 6 months on the study at MD Anderson Cancer diagnostic laboratory or Quest Diagnostics.

### Celecoxib treatment of Apc^Δ580^ mice

Mouse care and experimental protocols were approved and conducted in accordance with the guidelines of the Animal Care and Use Committee of MD Anderson. Apc^Δ580^-flox mice were bred with CDX2-cre mice that specifically expressed Cre recombinase in the colon to generate *Apc*^*Δ580*^-flox^(+/-)^; CDX2-*Cre* ^(+/-)^ mice (*Apc*^*Δ580(+*/-)^, designated as Apc^Δ580^ mice in this study), in which one allele has an *Apc* codon 580 frame-shift mutation in colons, as described previously (20).

Celecoxib powder was obtained from commercially available celecoxib capsules (200 mg/capsule, NDC 69097-421-07, Cipla USA, Inc.) and mixed with 4% carboxymethylcellulose sodium (cat. #C9481, Sigma) to create a celecoxib suspension at a concentration of 20 mg/ml. We orally administered the celecoxib suspension via gavage to 14-week-old Apc^Δ580^ mice with existing colon tumors (*n* = 6) (Supplementary Figure 1) at a final dose of 200 mg/kg/day for 5 consecutive days. The mice were euthanized, and the colonic tumors and adjacent normal colonic tissues were harvested separately and immediately flash frozen in liquid nitrogen. The tissue samples were transferred and stored in a −80°C freezer until they were analyzed.

**Figure 1.**
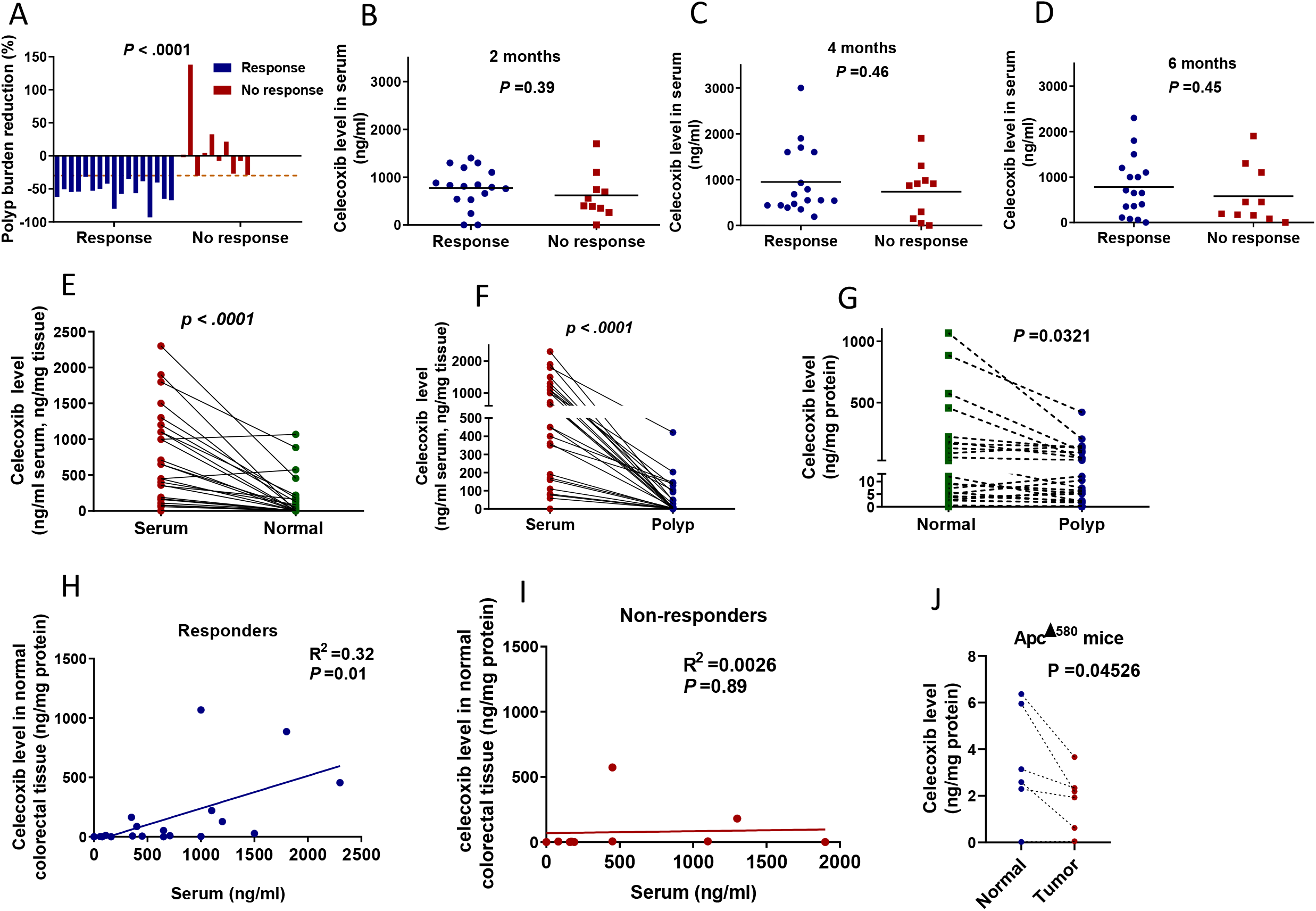
Celecoxib levels in colorectal tissues and sera in humans and mice A-I. FAP patients (*n* = 27) received 400 mg of oral celecoxib twice per day for 6 months. Colonoscopy was performed to assess polyp burden and collect colorectal tissue samples before and 6 months after celecoxib treatment. **(A)** Celecoxib response statuses for each patient, based on the response cut-off of polyp tumor burden reduction by > 30% from the baseline (brown dotted line). **(B-D)** Comparisons of celecoxib levels in sera at 2 **(B)**, 4 **(C)**, and 6 **(E)** months of celecoxib treatment in patients who did and did not experience a celecoxib response. Lines represent means. **(E and F)** Comparisons between celecoxib levels in paired sera and normal colorectal tissues **(E)** or polyps **(F). (G)** Comparison of celecoxib levels in paired normal colorectal tissues and polyps. **(H)** Correlation analyses of celecoxib levels between sera and normal colorectal tissues in the 17 of 27 FAP patients who experienced a response. **(I)** Correlation analyses of celecoxib levels between sera and normal colorectal tissues in the 10 of 27 FAP patients who did not experience a response. **(J)** Comparison of celecoxib levels in normal and polyp colorectal tissue samples from Apc^Δ580^ mice. We treated 14-week-old Apc^Δ580^ mice orally with celecoxib (200 mg/kg/day) via gavage for 5 consecutive days (*n* = 6 mice) and then euthanized. Celecoxib levels of the paired polyps and adenomas and their adjacent normal colorectal tissues were measured and shown. The results are presented as mean ± SEM.

### Liquid chromatography and tandem mass spectrometry measurements (LC/MS/MS) of celecoxib levels and LOX products

Lipid products and celecoxib were extracted in a manner similar to previously published methods (15). In brief, each frozen biopsy tissue sample was cut into approximately 1- to 2-mm strips. Samples were transferred to sealed microcentrifuge tubes, and 500 μl of ice-cold tissue homogenization buffer were added (21). The samples were homogenized by an Ultrasonic Processor (Misonix) at 0°C for 3.5 minutes twice, with a 1-minute rest, and then centrifuged at 10,000 rpm for 5 minutes at 4°C. A 400-μl aliquot of the supernatant was transferred to a glass tube; 600 μl of PBS buffer (containing 1 mmol/L EDTA and 1% butylated hydroxytoluene and 10 μl of 12-, or 15-hydroxyeicosatetraenoic acid (12-, or 15-HETE); leukotriene B_4_ [LTB_4_]; or 13-HODE; 1 μg/ml) was added, and samples were acidified with 0.5 N HCl (pH = 3.2 to 3.3). Lipid products were extracted by adding 2 mL of ethyl acetate and vertexing for 30 seconds, followed by centrifugation at 2000 rpm for 5 minutes at 4°C. The upper organic layer was collected, extraction was repeated two more times, and the organic phases from three extractions were pooled and evaporated to dryness on ice under a stream of nitrogen. Samples were reconstituted in 100 μl of methanol and ammonium acetate buffer (10 mmol/L at pH 8.5; 70:30, v/v) before liquid chromatography-tandem mass spectrometry (LC-MS/MS) analysis. The protein concentration was determined using a Bradford protein assay (Bio-Rad). LC-MS/MS analyses for eicosanoids were performed using a Quattro Ultima tandem mass spectrometer (Micromass) equipped with an Agilent HP 1100 binary-pump HPLC inlet, as described previously (22). The LC/MS/MS for celecoxib was operated under the similar condition as described above for eicosanoid analysis with minor changes. Briefly, ten microliters of the sample were injected on a Luna 3 um phenyl-hexyl 2×150 mm analytical column (Phenomenex). Celecoxib was detected and quantified by operating the mass spectrometer in electrospray negative ion mode and monitoring the transition *m/z* 380.2>316.1. The results are presented as ng/mg protein.

For the serum celecoxib, an aliquot (100 μl) of serum was diluted with an equal volume of 10 mM ammonium acetate, pH 8.5. To this solution, 4 ml of hexane: ethyl acetate (1:1, v/v) was added; the mixture was vortex mixed for 5 min. and then centrifuged at 4000 rpm at 5°C for 5 min. The extraction was repeated twice, and the upper organic layer was collected, pooled and evaporated to dryness under a stream of nitrogen at room temperature. The sample was then reconstituted in 200 μL of methanol: 10 mM ammonium acetate, pH 8.5 (1/1. v/v). The celecoxib level in the samples was determined by LC/MS/MS. Quantification of serum celecoxib was done by comparing the sample peak areas to a standard curve constructed from peak areas of extracted serum sample spiked with known amounts of celecoxib. The results are presented ng/ml.

### Nutrient intake

We measured the intake of nutritional, mineral, and vitamin supplements and medications. Dietary intake and alcohol intake were assessed using a 137-item semi-quantitative food frequency questionnaire (FFQ) that elicited usual intake over the previous 6 months. Martinez et al. described this FFQ and its measurement characteristics (23). FFQ data were entered into the Food Frequency Data Entry and Analysis Program; 49 macronutrients and micronutrients, as well as individual fatty acids, were analyzed using nutrient and gram weight information from the Food Intake Analysis System USDA Survey Nutrient Data Base (U.S. Department of Agriculture, Agricultural Research Service, 1997; ON: Nutrient Database for Individual Intake Surveys; 1994-1996 Continuing Survey of Food Intakes by Individuals, and 1994-1996 Diet and Health Knowledge Survey [CD-ROM]). Intake of hormone replacement therapy, vitamins, and other nutritional supplements was measured using a medication questionnaire.

### Statistical methods

For paired two-group comparisons (e.g., normal tissue and polyp from the same subject), we used a paired *t*-test. Analysis of variance (one-way) with Bonferroni adjustments was used for multiple group comparisons. All tests were two-sided, and significance was defined as *P* < 0.05. Data were analyzed using GraphPad Prism software 8.0.0 (GraphPad Software).

## RESULTS

### Patient study sample

Of the 47 patients enrolled in the study, 27 completed the procedures required for the reported laboratory analyses and were included in the final population. Patients’ demographic and clinical characteristics and treatment schedules and the polyp burden assessments used to determine response were described previously (19).

### Dietary intake and clinical response

Celecoxib responders were defined as those who experienced a reduction in polyp tumor burden of more than 30% of the baseline (19) (**Figure 1A**). Out of 27 FAP patients included in this study, 17 are responders, and 10 are non-responders. Responders and non-responders were similar in regard to age, sex, and BMI (**Table 1**). Dietary intake, as measured by FFQ, did not significantly differ in terms of the intake of calories and various dietary components, including total fat, arachidonic and linoleic acids between responders and non-responders (**Table 1**).

**Table 1.** Comparison of demographic characteristics and mean serum levels of nutrients between responders and non-responders

| **Variable** | **Responders (n = 17)** | **Non-responders (n = 10)** | **P value** |
| --- | --- | --- | --- |
| Median age, years (25%-75%) | 32 (18.75-44.5) | 31 (22.5-47.5) | 0.75 |
| Sex (%) |  |  |  |
| Male | 50 | 59% | 0.71 |
| Female | 50 | 41% |  |
| Median BMI (25%-75%) | 26.7 (22.28-30.8) | 26.34 (22.5-31.24) | 0.87 |
| Energy intake | 1651 | 1908 | 0.481543 |
| Protein | 65.44 | 72.78 | 0.565846 |
| Carbohydrate | 196.3 | 238.3 | 0.369143 |
| Total fat | 67.72 | 76.59 | 0.620910 |
| Saturated fat | 22.69 | 27.56 | 0.454275 |
| Monounsaturated fat | 24.78 | 28.03 | 0.583000 |
| Polyunsaturated fat | 15.31 | 15.26 | 0.991848 |
| Cholesterol | 184.0 | 223.8 | 0.397686 |
| Dietary fiber | 15.43 | 16.61 | 0.772242 |
| Vitamin A IU | 23715 | 9258 | 0.479369 |
| Vitamin A RE | 2641 | 1189 | 0.479977 |
| Carotene RE | 2236 | 795.0 | 0.479850 |
| Thiamine | 1.271 | 1.405 | 0.545762 |
| Riboflavin | 1.687 | 1.674 | 0.973229 |
| Niacin | 16.64 | 18.41 | 0.539613 |
| Vitamin B6 | 1.537 | 1.491 | 0.879802 |
| Folate | 333.0 | 381.5 | 0.450087 |
| Vitamin B12 | 3.822 | 4.023 | 0.827999 |
| Vitamin C | 108.0 | 122.1 | 0.675367 |
| Vitamin E | 9.007 | 8.658 | 0.870050 |
| Calcium | 902.1 | 876.9 | 0.925326 |
| Phosphorous | 1143 | 1208 | 0.788561 |
| Magnesium | 245.7 | 266.9 | 0.652355 |
| Iron | 12.56 | 13.90 | 0.563903 |
| Zinc | 10.43 | 10.92 | 0.810278 |
| Copper | 1.072 | 1.192 | 0.589273 |
| Sodium | 2640 | 2953 | 0.591152 |
| Potassium | 2542 | 2574 | 0.952888 |
| Alcohol | 3.422 | 1.335 | 0.184077 |
| Moisture | 1548 | 1605 | 0.898905 |
| Butyric | 0.4410 | 0.5317 | 0.635469 |
| Caproic | 0.2360 | 0.2884 | 0.610695 |
| Caprylic | 0.2051 | 0.2951 | 0.273041 |
| Capric | 0.3763 | 0.4971 | 0.407208 |
| Lauric | 0.7896 | 1.233 | 0.168766 |
| Myristic | 1.980 | 2.531 | 0.439120 |
| Palmitic | 12.20 | 14.43 | 0.511914 |
| Stearic | 5.783 | 7.003 | 0.415536 |
| Palmitoleic | 1.158 | 1.444 | 0.374002 |
| Oleic | 23.01 | 25.86 | 0.603110 |
| Gadoleic | 0.1077 | 0.1209 | 0.580432 |
| Erucic | 0.01710 | 0.02730 | 0.141708 |
| Linoleic | 13.57 | 13.45 | 0.977465 |
| Linolenic | 1.389 | 1.362 | 0.954841 |
| Parinaric | 0.001487 | 0.002957 | 0.088584 |
| Arachidonic | 0.09946 | 0.1198 | 0.432583 |
| EPA | 0.01193 | 0.02794 | 0.019649 |
| DPA | 0.01030 | 0.01661 | 0.083952 |
| DHA | 0.03470 | 0.06625 | 0.048061 |
| Caffeine | 86.01 | 90.56 | 0.913827 |

|  |  |  |  |
| --- | --- | --- | --- |
| Theobromine | 29.32 | 43.01 | 0.248597 |
| Selenium | 76.66 | 88.24 | 0.435442 |

### Celecoxib levels in sera and colorectal tissues and clinical response

We first determined whether celecoxib bioavailability affected clinical response by measuring celecoxib levels in sera and colorectal tissues. Celecoxib levels in sera were similar at 2, 4, and 6 months of celecoxib treatment between responders and non-responders (**Figure 1B-D**). Celecoxib levels were significantly lower in normal and polyp colorectal tissues than in their paired serum levels (**Figure 1E and F**). Celecoxib levels (mean ± SEM) were significantly lower in polyps (48.99 ± 16.89 ng/mg protein) than in normal colorectal tissues (134.70 ± 50.51) (**Figure 1G**). Celecoxib levels in normal colorectal tissues were significantly correlated with those in sera in the 17 responders (**Figure 1H**) but not in the 10 non-responders (**Figure 1I**). Celecoxib levels were also significantly lower in colorectal tumors (1.80 ± 0.52 ng/mg protein) than in paired normal colorectal tissues (3.40 ± 0.99) in celecoxib-treated Apc^Δ580^ mice (**Figure 1J and Supplementary Figure 1**).

### Levels of LOX oxidative products and celecoxib clinical response in FAP patients

We assessed the effects of celecoxib on the oxidative metabolism of LOX linoleic and arachidonic acids by measuring the major LOX oxidative products: 13-HODE, 15-HETE, 12-HETE, and LTB4 levels in colorectal tissues in 27 FAP patients. 13-HODE levels were higher than 15-HETE, 12-HETE, and LTB4 levels in both normal colorectal tissues and polyps (**Supplementary Figures 2A and B**) and in normal colorectal tissues than in polyps (23.03 ± 3.57 versus 14.56 ± 1.95 ng/mg protein, **Figure 2A**), while 15-HETE and 12-HETE levels were similar between normal and polyps (**Figure 2D and G**). LTB4 levels were also similar between normal and polyps (median = 0.07 for normal tissue and 0.065 ng/mg protein for polyps) and low and undetectable in 55% of polyps and 50% of normal mucosa (**Figure 2J**).

**Figure 2.**
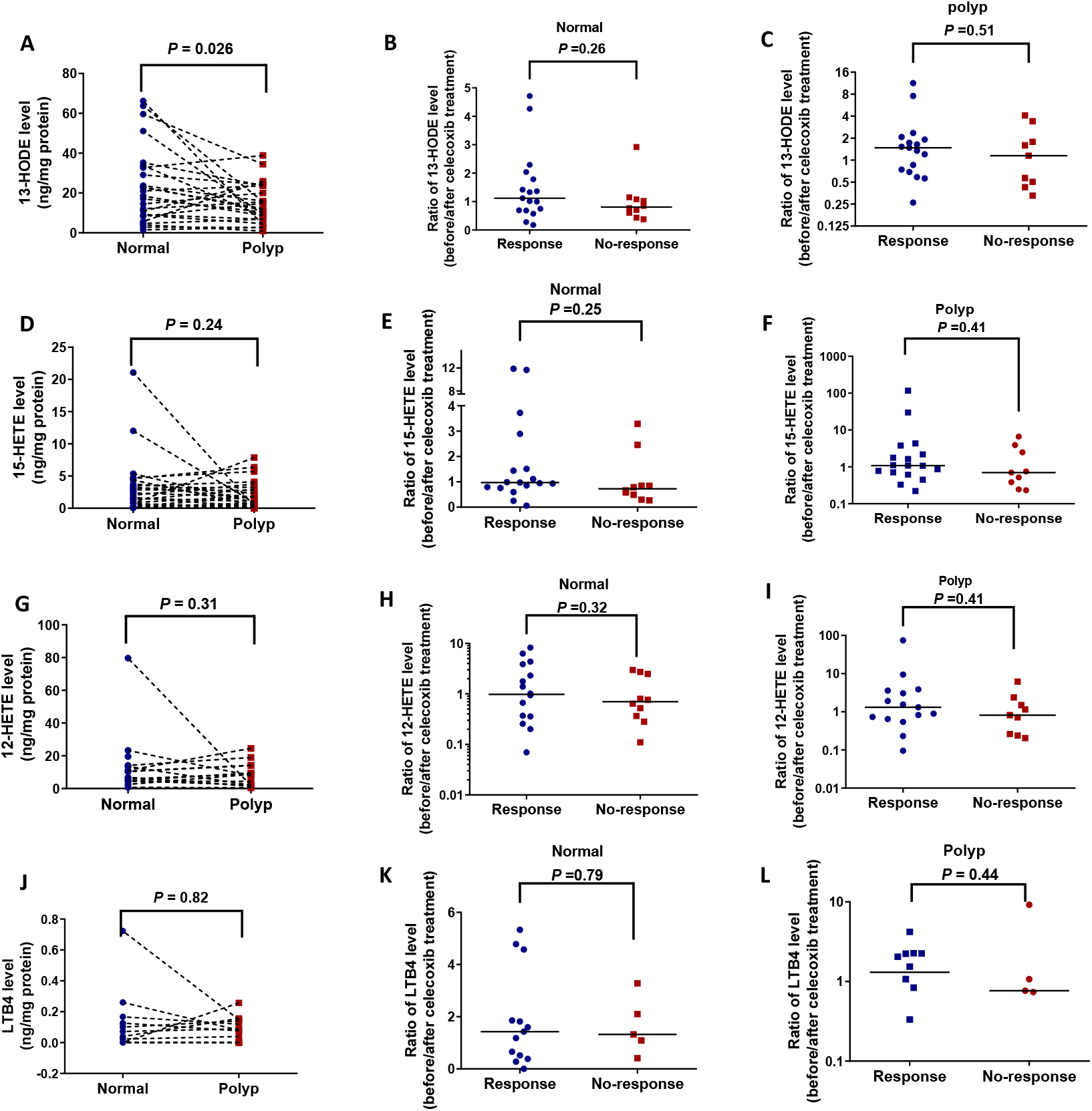
Effects of celecoxib on generations of colorectal lipoxygenase products in FAP patients. The indicated lipoxygenase products were measured by LC/MS/MS in normal tissues and polyps before and 6 months after celecoxib treatment (*n* = 27). (**A, D, G, and J**) Comparisons of 13-HODE **(A)**, 15-HETE **(B)**, 12-HETE **(G)**, and LTB_4_ **(J)** levels between paired normal and polyp colorectal tissue samples from FAP patients prior to celecoxib treatment (*n* = 27). **(B-L)** Comparisons of 13-HODE (**B** and **C**), 15-HETE (**E** and **F**), 12-HETE (**H** and **I**) and LTB4 (**K** and **L**) ratios of after over before treatment in normal colorectal tissues and polyps in patients who did (*n* = 17) and did not (*n* = 10) experience a celecoxib response. Lines represent medians.

The ratios of 13-HODE levels of after over before celecoxib treatment trended to be higher in normal colorectal tissues from 17 responders (mean ± SD: 1.50 ± 1.26) than in those from non-responders (0.99 ± 0. 73) (*P* = 0.26, **Figure 2B**); we also found no statistically significant difference between the two groups in polyps (*P* = 0.51; **Figure 2C**). The ratios of 15-HETE (**Figure 2E and F**), 12-HETE (**Figure 2H and I**), and LTB4 (**Figure 2K and L**) levels of after over before celecoxib treatment were not statistically different in normal colorectal tissues and polyps between the 17 patients who experienced a response and the 10 patients who did not experience a response.

## Discussion

Our study is the first to evaluate the bioavailability of celecoxib and byproducts of the lipoxygenase pathway in sera, colorectal normal tissues, and polyps from patients with FAP. We found that celecoxib levels were significantly lower in colorectal polyps and tumors than in their paired normal-appearing tissues in humans and mice with Apc^580^ mutation; a correlation was found between celecoxib levels in sera and normal colorectal tissues in celecoxib responders, but not in celecoxib non-responders.

Celecoxib bioavailability was reduced in colorectal polyps. Both non-responders and responders had good celecoxib systemic bioavailability, as measured by serum levels 2, 4, and 6 months after celecoxib treatment. However, celecoxib levels were significantly lower in polyps than in paired normal colorectal tissues in FAP patients. The lack of correlation of celecoxib levels between sera and normal colorectal tissues in non-responders further suggests that reduced bioavailability in colorectal tissues contributed to the failure of celecoxib to prevent colorectal tumorigenesis. Of note, normal colorectal tissue levels of celecoxib in 70% of non-responders were < 15% of their paired celecoxib levels in sera. Studies in Apc^Δ580^ mice that experimentally model human FAP similarly showed that celecoxib levels were lower in colorectal tumors than in their paired normal-appearing colorectal tissues. This observation further supports the concept that the reduction of celecoxib bioavailability limits the chemo-preventive effectiveness of celecoxib.

The differentially reduced bioavailability of celecoxib in colorectal polyps may be the result of differential vascular delivery or drug metabolism between normal and polyp tissues. In advanced cancer, reduced drug bioavailability in cancer tissues has been well described and documented to limit anti-cancer treatment activity (24) (25). Whether this principle applies to earlier stages of tumorigenesis is poorly understood. To our knowledge, the colorectal tissue bioavailability of celecoxib has not been reported previously. Our data demonstrated the differential celecoxib bioavailability in colorectal polyps and its possible effects on celecoxib’s chemo-preventive activity. Thus, our data support the need to monitor celecoxib levels in tissues rather than in sera to better predict chemo-preventive response. Further studies are needed to assess the potential mechanisms of reduced celecoxib bioavailability to identify approaches to enhance chemo-preventive activity.

Our results confirmed that the reduction of 13-HODE in colorectal tumorigenesis is differential compared to other LOX products (13) (15) and unrelated to the difference in the intake of its substrate (i.e. linoleic acid) or other dietary factors. 13-HODE levels in normal colorectal tissues trended to be higher in celecoxib responders compared to those in non-responders, however, the difference was statistically non-significant. Our inability to detect a statistically difference in celecoxib modulation of 13-HODE in celecoxib responders of FAP patients could be related to: 1. the small sample size (17 subjects); and 2. low levels of celecoxib in colorectal tissues at the time of the measurements; 9/17 subjects in this responder group had celecoxib levels of less than 50 ng/mg (approximately 0.13 µM) protein in normal colorectal tissues. These celecoxib levels are significantly below levels that are required (12.5-40 µM) to induce 15-LOX-1 expression and increase 13-HODE levels in preclinical studies(14,18). These findings underscore the need to measure anti-tumorigenic drugs tissues concentration when evaluating the effects of these agents on putative biomarkers to ensure that the modulation of these biomarkers is indeed linked to the studied drugs.

The major limitation of our study is its relatively small size, which is reflective of the difficulty of conducting studies in FAP patients, secondary to the relative rarity of this syndrome. Despite our small sample size, we were able to detect significant celecoxib reduced bioavailability in colorectal polyps, and a correlation of celecoxib levels between sera and colorectal tissues in patients who experienced a response. Further studies are needed to validate these findings and assess the utility of measuring the levels of chemopreventive agents in normal colorectal and tumor tissues as potential biomarkers for response.

In conclusion, our findings demonstrate the need to consider the bioavailability of chemo-preventive agents as a potential rate-limiting factor to their effectiveness and the need to identify strategies to discover and modify this potentially important resistance mechanism.

## Data Availability

All data referred to in the manuscript are available by requested.

## Supplemental information

Supplemental Information can be found online.

## Acknowledgments

This work was supported by the National Cancer Institute (R01-CA106577 and R01-CA137213 to I.S., R01-CA144053 to P.Y, and R03-CA235106 to X. Z). This study made use of the MD Anderson Cancer Center Genetically Engineered Mouse Facility. We acknowledge all our lab team members, patients, and patients’ family members who contributed their time and valuable resources to this research. We thank Ms. Ann M. Sutton at Scientific Publications at MD Anderson Cancer Center for editing the manuscript.

## Author contributions

I.S. conceived the study and designed the experiments. S.A. and J.M. collected human colorectal tissues and blood samples, X.Z. conducted mouse experiments, and B.W. performed liquid chromatography– tandem mass spectrometry to measure LOX oxidative products and celecoxib levels and P.Y. evaluated the results. S.A assisted with nutritional data analyses and contributed to the manuscript drafting. I.S, P.Y. and X.Z. analyzed the data. I.S., P.Y. and X.Z. wrote the manuscript. R.S.D guided the nutritional analyses and provided conceptual feedback for the manuscript.

