## Supplemental Figures 1 and 2 for "Celecoxib Colorectal Bioavailability and Chemopreventive Response in Familial Adenomatous Polyposis Patients"

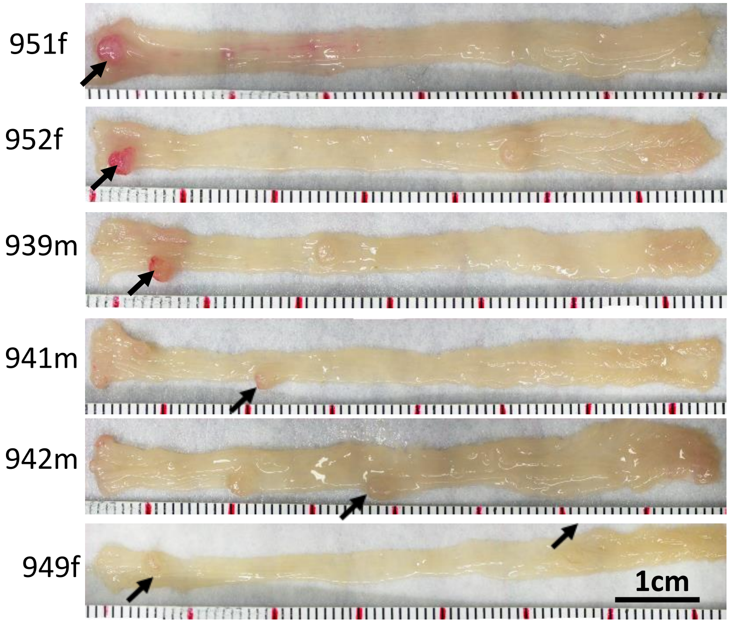


**Supplementary Figure 1. Colorectal tumor formation in Apc$^{\Delta580}$ mice.** 14-week-old Apc$^{\Delta580}$ mice were orally treated via gavage with the celecoxib suspension at a final dose of 200 mg/kg/day for 5 consecutive days ($n$ = 6 mice). Mice were killed and colorectal images were captured and shown (**A**). Black arrows show the colonic tumors.

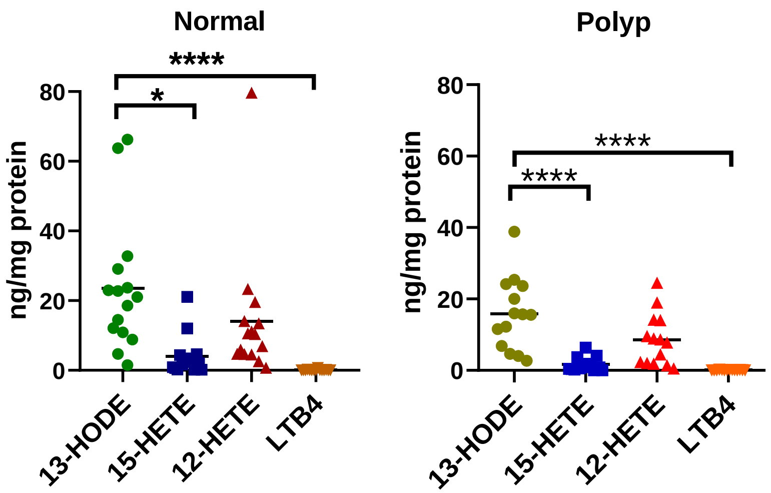


**Supplementary Figure 2. Levels of 13-HODE, 15-HETE, 12-HETE, and LTB4 in normal and polyp colorectal tissues**. The colorectal tissues were obtained from FAP patients prior to celecoxib treatment, and those indicated lipoxygenase products were measured by liquid chromatography/tandem mass spectrometry ($n$ = 27). The data are presented as mean ± SEM. **P < .05* and ***** P < .0001*; two-sided one-way ANOVA.
